# Developing a Discrete Choice Experiment to Understand Patient Preferences in Resource-Limited Settings: a Six-Step Guide

**DOI:** 10.1101/2020.10.05.20206557

**Authors:** Sarah Moor, Andrew K. Tusubira, Ann R Akiteng, Evelyn Hsieh, Christine Ngaruiya, Tracy L. Rabin, Nicola L. Hawley, Mari Armstrong-Hough, Rachel Nugent, Robert Kalyesubula, Christine Nalwadda, Isaac Ssinabulya, Jeremy I. Schwartz

## Abstract

A discrete choice experiment (DCE) is a method to quantify preferences for goods and services in a population. Participants are asked to choose between sets of 2 hypothetical scenarios that differ in terms of particular characteristics. Their selections reveal the relative importance of each “attribute”, or characteristic, and the extent to which people consider trade-offs between characteristics. DCEs are increasingly used in healthcare and public health settings as they can inform the design of health-related interventions to achieve maximum impact. Specific efforts must be made in the development process to ensure relevance of DCEs to the communities in which they are administered. Herein, we build upon gaps in the prior literature by offering researchers a step-by-step process to guide DCE development for resource-limited settings, including detailed methodological considerations for each step and a specific actionable approach that we hope will simplify the process for other researchers. We present the 6 steps we followed to develop a DCE to evaluate patient preferences for management of hypertension and diabetes in rural Uganda. These steps are: 1) formative work; 2) attribute selection; 3) attribute level selection; 4) DCE design selection; 5) determination of attribute level combinations; and 6) assessment and enhancement of tool comprehensibility. We describe each of these steps in detail to ease the development process for researchers looking to develop locally contextualized, end-user-centric health interventions.

## Introduction

Discrete choice experiments (DCEs) are quantitative tools used to understand decision-making and elicit preferences. Participants are asked to choose between hypothetical scenarios that describe goods or services. Each scenario is described by characteristics, called *attributes*, with each assigned a specific *attribute level*. For example, the distance required to travel to purchase a good or service is an attribute. Attribute levels are the value of distances one could travel.^1 2^ Participants are presented multiple pairs of scenarios, called *choice sets*, and are asked to select which scenario they would prefer. Subsequent choice sets vary the combination of attribute levels. Selection of various scenarios reveals the relative impact of each attribute on participant decision-making and the extent to which individuals are willing to trade an attribute for another.^2^

DCEs are increasingly being used within the health field to understand patient preferences, including assessment of preferences for migraine treatment, vaccinations, and HIV testing and treatment.^3-8^ In resource-limited settings (RLS), eliciting healthcare preferences is particularly important in order to appropriately allocate resources to achieve the greatest impact. When designing a DCE for use in RLS, additional factors must be considered, such as health literacy and education levels, which are likely to be heterogeneous among the population.^9^ Most published DCE studies focus on the results rather than how the DCE was generated. However, there are numerous methodological considerations that researchers must address along the way and these are under-developed in the existing literature, including in the standard DCE textbook in which such considerations of setting and participant characteristics are not discussed.^10^

Our DCE was part of a research project in which we sought to understand how patients with hypertension and diabetes in rural Uganda place relative value on various aspects of their chronic care provision. Our goal was to use these findings to guide the development of locally relevant health system interventions. In beginning our work, we used The Conjoint Analysis Applications in Health Checklist (Figure 1) which outlines the fundamentals of creating a valid DCE as well as the paper by Mangham *et al* which outlines the various unique factors to consider when designing a DCE for use in low-income countries.^9 11^ While this prior literature offers guiding principles and decisions to be addressed in DCE development, it lacks detail of how to make each of the decisions or explanation of the benefits and drawbacks of the available options. Herein, we build upon this literature by presenting a step-by-step process for DCE development with an RLS focus. We offer detailed methodological considerations for each step and a specific actionable approach that we hope will simplify the process for other researchers seeking to develop DCEs for healthcare research in RLS.

**Figure 1:**
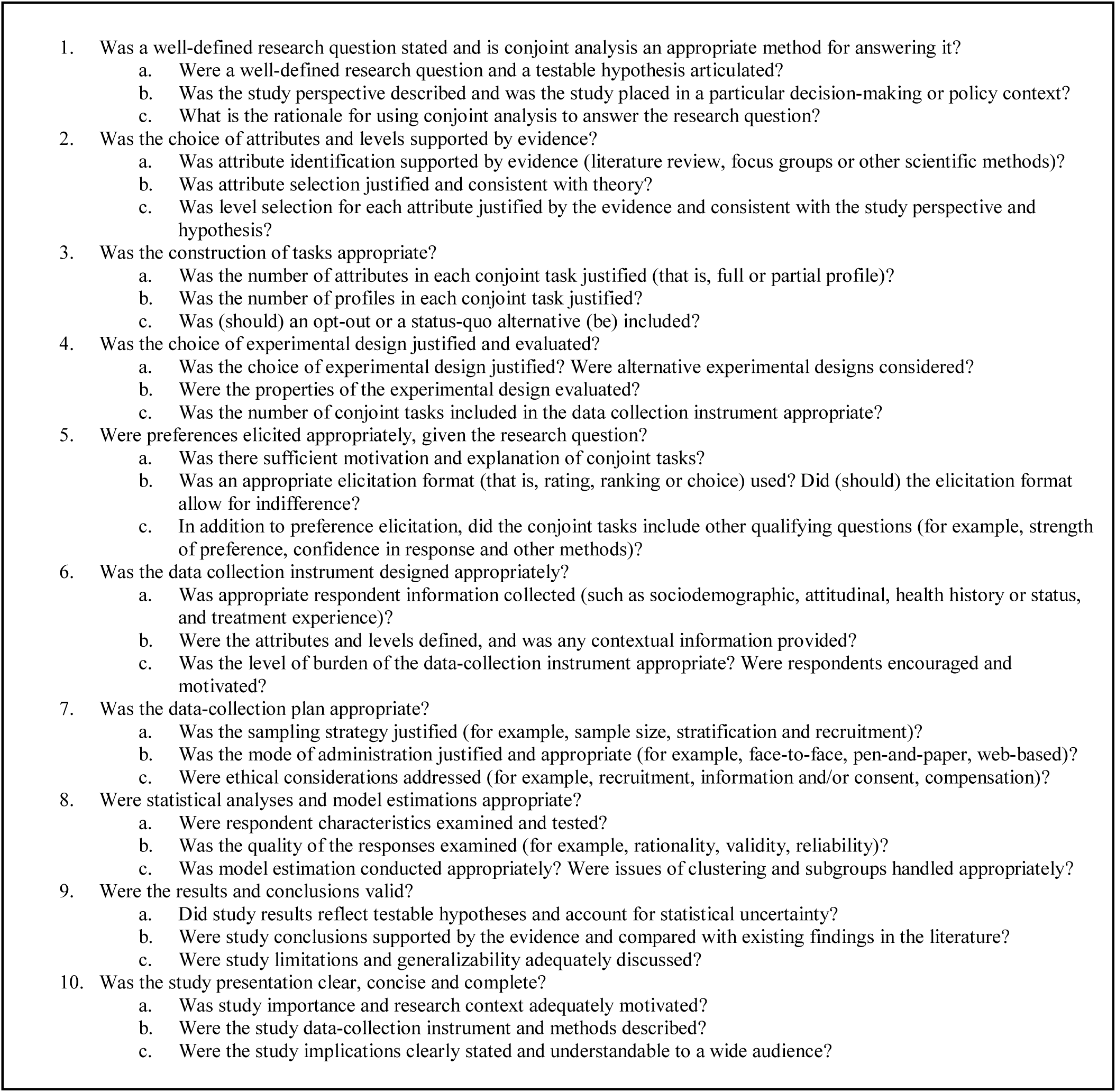
Conjoint Analysis Applications in Healthcare Checklist adapted from [Bridges, J.F., Hauber, A.B., Marshall, D., Lloyd, A., Prosser, L.A., Regier, D.A., Johnson, F.R., and Mauskopf, J. (2011). Conjoint analysis applications in health--a checklist: a report of the ISPOR Good Research Practices for Conjoint Analysis Task Force. *Value Health* 14, 403-413.] (Bridges et al., 2011)

### The 6-Step Process

The 6 steps we followed in developing our DCE included: formative work, attribute selection, attribute level selection, DCE design selection, determination of attribute level combinations, and assessment and enhancement of tool comprehensibility. Figure 2 illustrates these steps and the specific tasks to be completed within each step. Careful consideration throughout progression of this workflow is crucial to generating a locally-relevant, rationally-designed DCE.

**Figure 2:**
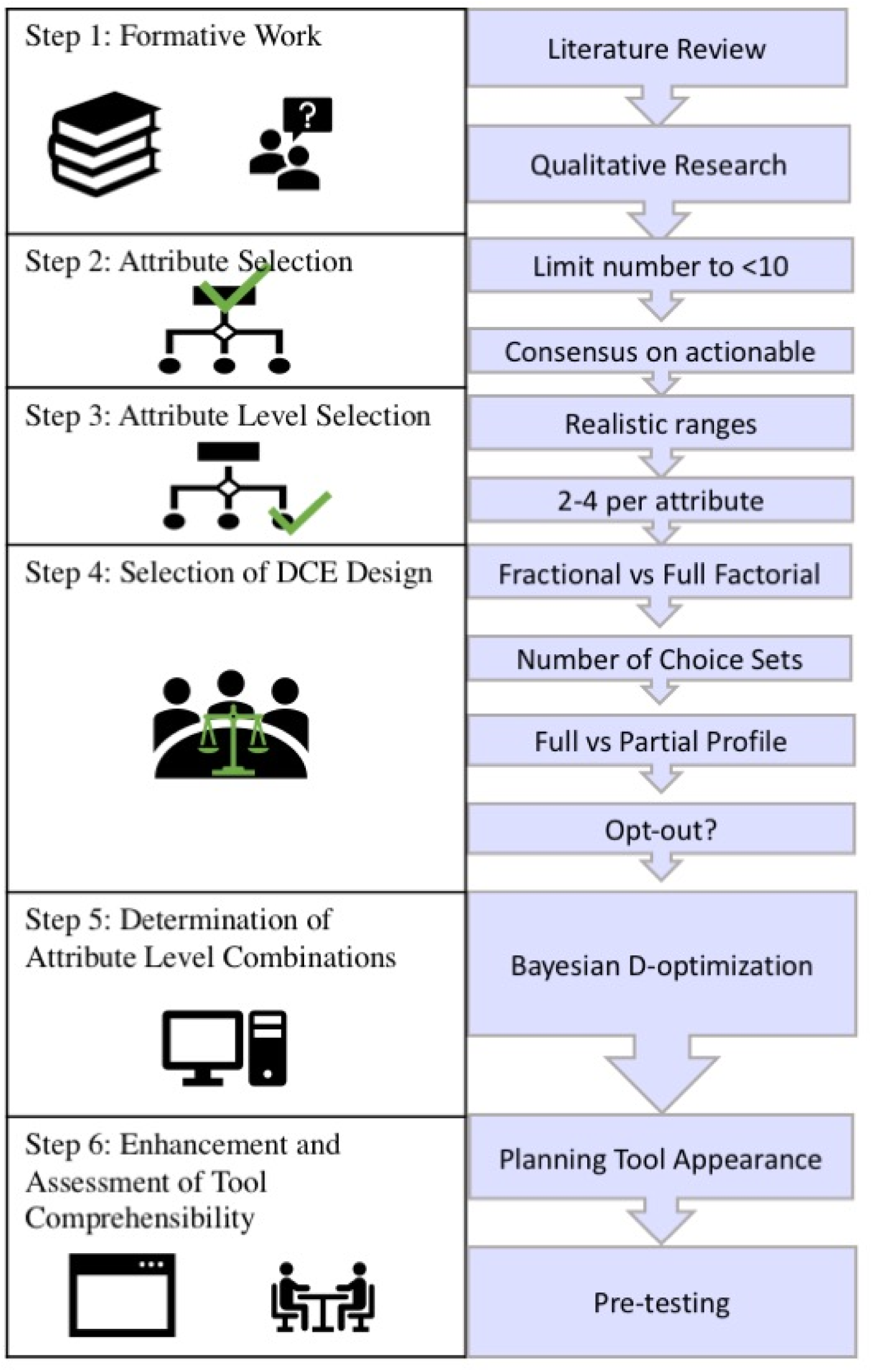
Six-step process for the development of a locally-relevant and rationally designed DCE.

#### Step 1: Formative Work

We began by consulting the literature and conducting qualitative research. The literature review focused on previous applications of DCEs in low- and middle-income countries (LMIC). We searched PubMed with the terms “discrete choice” and “low- and middle-income countries.” This search yielded 56 results which directed the qualitative interview guide. For instance, Zanolini et al. found that provider attitude is a significant driver of preferences in patients with HIV in Zambia.^12^ To evaluate whether provider attitude also influences patients with non-communicable diseases (NCDs) in rural Uganda, we included questions in our in-depth interview guide about relationships with healthcare providers. This example illustrates how the literature review guided our participant interviews.

We interviewed 18 participants in Nakaseke District, Uganda with hypertension, diabetes, or both about their experiences accessing healthcare and medicines for these conditions. We chose this sample size as it would likely achieve thematic saturation.^13^ Participants were asked about 3 themes: accessing care, accessing medicines, and relationships with healthcare providers. Interviews were conducted in Luganda by a single research assistant, then transcribed and translated into English. A directed coding approach was used to code the transcripts and identify attributes and their appropriate levels to be included in the DCE. Directed coding analysis is performed when there is a pre-existing direction about what the researcher is looking for in the qualitative data.^14^ This approach allowed 3 qualitative coders, including 2 Ugandan coders, to identify concepts in the data that could be attributes in the DCE. Consensus was reached between coders through an iterative coding process and continuous discussion to ensure agreement on code assignments and definitions. 10 tentative attributes emerged: the presence of peer support groups, health education, interactions with healthcare providers, range of treatments, perceived quality of care, waiting time, use of herbal medicine, getting to the facility, costs of treatment, and availability of medicines.

#### Step 2: Attribute Selection

It is important to limit the number of attributes to fewer than 10 to minimize participant fatigue during the completion of the task.^9^ DCE tools that present too significant a cognitive burden are likely to lead to respondents using a simplifying heuristic to make decisions, such as selecting based on a single attribute.^11^ Previous DCEs administered in LMICs incorporated 5 or 6 attributes, which led us to select 6 attributes.^12 15 16^ To determine which of the 10 tentative attributes would be included in the DCE, a multi-disciplinary team of experts in public health, medicine, anthropology, and NCDs from Makerere University, Yale University, and New York University gathered to discuss the tentative attributes. The Nominal Group Technique was used to reach a consensus on which attributes would be included in the final DCE tool.^17^ Using this well-established method, the experts were presented the tentative attributes, discussed the pros and cons of including each in the DCE, and rank-ordered each attribute. The rank order was discussed and a final re-ranking was performed. Primary reasons for the inclusion of attributes were amenability to intervention development and impact on patients’ experience of their care based on interview results. The team reached consensus on a list of 6 attributes: presence of peer support groups, education at the facility, interactions with healthcare providers, getting to the facility, costs of treatment, and availability of medicines. We excluded the use of herbal medicines, range of treatments, perceived quality of care, and waiting time because they were felt to be represented by 1 of the other 6 priority attributes, unamenable to intervention or ambiguous in interpretation based on the interview data.

#### Step 3: Attribute Level Selection

To determine levels for each attribute, the team considered the range of situations encountered by patients as described in the in-depth interviews. In the attribute levels, we sought to reflect the realistic range of situations that patients currently encounter.^9^ For instance, the “costs of treatment” attribute presents a range of monthly costs, between zero and 20,000 Ugandan Shillings (approximately 5 USD). Free treatment represents patients attending the public district hospital for appointments and medicines. Each of the other attribute levels presented reflects the reported out of pocket costs when patients attend private clinics and/or pharmacies. To ensure that we presented a realistic range of attribute levels for each attribute, we reviewed the ranges as reported in interviews and verified these with members of the team who are healthcare workers in the district where the research was based. Researchers might choose to include potential attribute levels that describe aspirational, rather than currently realistic, healthcare options. This was the case for our attribute level “I receive more than a month’s worth of medicine from the facility pharmacy” within the “availability of medicines for my condition” attribute. Though not widely offered, dispensing of greater than 1 month’s supply of medicines is an approach increasingly used in differentiated service delivery for HIV care in LMICs.^18^ While this option is not currently available for patients with NCDs in Uganda, we included it as an attribute level because of the research team’s interest in future expansion of differentiated service delivery models into NCD care in Uganda. The final attributes and attribute levels we determined based on this process are shown in Figure 3. Consistent with the Conjoint Analysis Good Practice Checklist, we avoided presenting attribute levels as ranges. Ranges result in variability in how respondents interpret each level, thereby limiting analysis. Additionally, we limited each attribute to a maximum of 4 attribute levels in accordance with good practice.^11^

**Figure 3:**
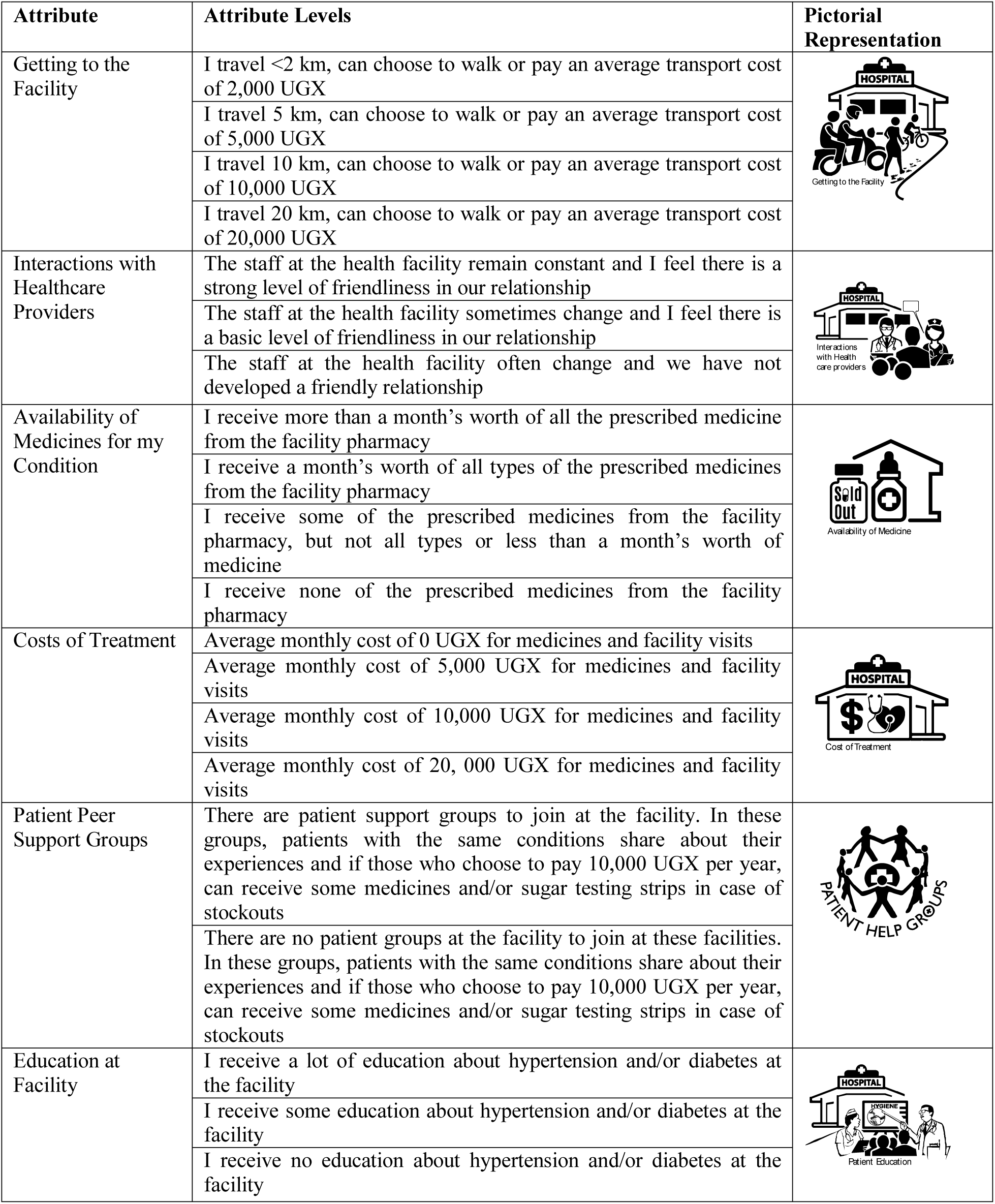
Attribute and their respective levels included in the discrete choice experiment. (1 USD is approximately equal to 3700 Ugandan Shillings)

Determining how patients think about a given attribute was an important consideration in our discussion of attributes. We aimed to define each attribute level using terms that patients used in the interviews. In particular, the attribute “getting to the facility” was challenging to frame due to the diverse ways in which patients described travel in the context of seeking healthcare. Patients typically described either the cost of transport or the length of the journey as the challenge in getting to the healthcare facility. In an attempt to capture both types of descriptions, we created attribute levels reflecting both distance and cost. For instance, traveling 5 km by motorcycle typically costs about 5,000 Ugandan Shillings, so these were combined into an attribute level. We recognize this approach has drawbacks as we could not parse out whether participants’ selections were based on distance or cost.

#### Step 4: Selection of DCE Design

Health researchers who aim to design a DCE face a series of decisions in selecting the most appropriate design for their purpose. First, researchers must select between a *full* and a *fractional factorial design*. A full factorial design includes choice sets of all possible attribute level combinations for participants to choose between. Conversely, a fractional factorial DCE design presents only a subset of the possible combinations.^2^ Most DCEs, including ours, employ a fractional factorial design as the total number of possible combinations is too many to present to respondents.^12^

Second, we considered how many choice sets to present to each respondent. This DCE required respondents to consider 6 attributes in their selection of an ideal health facility for NCD care. The cognitive burden of this decision-making task could be compounded by variable health literacy levels in rural Uganda. The literature suggests that presenting between 8 and 16 choice sets is typically appropriate, but that reducing the number of choice sets also reduces the cognitive burden of the decision-making task.^9 11^ Following a discussion by the research team, we decided to present 8 choice sets to each participant to minimize the cognitive burden of the task. Additionally, as we did, pictorial representations of each attribute can be used to support the cognitive process of participants, especially in low literacy populations.

Third, in order to maximize the number of attribute level combinations included in the DCE, we created 3 different versions. Creating multiple versions of the tool is a strategy to increase the number of combinations presented to respondents, thereby increasing the coverage of hypothetical health scenarios evaluated.^12^ This resulted in a total of 24 choice sets, with each participant only completing 8 choice sets. When deploying the DCE, we block randomized distribution of the survey versions by study site to ensure adequate representation from each study site across all 24 choice sets.

Fourth, one must consider whether to create a DCE with a *full* or *partial profile*.^11^ A full profile design describes a DCE in which the attribute level presented for each attribute varies with every choice set presented.^19^ A partial profile describes a DCE in which certain attribute levels are omitted between choice sets and are used to handle DCEs with many attributes.^19^ In partial profile DCEs, respondents are instructed to assume that any omitted attributes are the same between the 2 options. However, we cannot know the attribute level that a respondent assumes for an omitted attribute, thereby limiting the ability of the DCE to reveal interactions between attributes.^19^ Since partial profile designs yield insufficient information to fully assess statistical interactions among attributes, it is preferable to use a full profile if the DCE has few attributes.

Finally, one must decide whether to include an *opt-out option*, in which participants are allowed to choose neither of the presented scenarios. The alternative is a *forced decision design*.^11^ While presenting respondents with an opt-out choice more realistically reflects real-world circumstances, it can limit data interpretation because it does not reveal any information about the preference between 2 options.^11^ It has also been demonstrated that participants with lower educational levels are more likely to opt-out of a decision task.^20^ A dual response design consists of choice sets in which the participant is forced to make a decision, then for that same choice set asked if they would prefer to opt-out. This design provides an efficient means to minimize the learning effect, which describes how participants are less likely to opt-out for choice sets that are presented later on in the choice experiment.^20^ However, this design does increase the complexity and time it takes participants to complete the task. Given the cognitively demanding nature of DCEs and our concern about a high level of non-response due to the limited education level of our target population, we used a forced decision design. Our final DCE consisted of a fractional factorial, full-profile forced decision design that presented 8 choice sets to each respondent.

#### Step 5: Determination of Attribute Level Combinations

The next step was optimizing the combination of attribute levels to improve the quality of the DCE output. We employed a Bayesian approach which takes into consideration common-sense assumptions about preferences within each attribute.^21^ For all variables, we assumed that the least burden on the patient would be the preferred option. For instance, for “costs of treatment” we assumed that paying less would be preferred, such that zero Uganda Shillings and 20,000 Ugandan Shillings (approximately 5 USD) monthly would be the most and least preferred options, respectively. This approach ensures that there are no uninformed choice sets presented and avoids creating a choice set in which a scenario is preferable in terms of all attributes.^21^ We used JMP software (Version 14) because we found it to have a user-friendly interface, an extensive user guide, and online user forum.^22^ JMP creates a Bayesian D-optimal DCE design in the multinomial logit model. D-optimality, in this case, means that the algorithm begins with a random starting design and over the course of the user’s determined number of random starts, selects the best design.^21^ Optimizing the combination of attribute levels helps create a DCE that yields more informative results.

#### Step 6: Enhancement and Assessment of Tool Comprehensibility

In step 6, we planned how to increase the comprehensibility of the tool upon its administration. We identified 3 domains that impact the comprehensibility of the tool: the introduction to, delivery of, and appearance of the instrument. In the written introduction to the tool displayed before the decision-making task, we provided an explanation of the task and a description of each of the attributes. Then, we planned for the tool to be delivered on tablets in the pre-testing and administration phases. Tablets aid understanding of the tool by allowing respondents to visually follow along with the research assistant’s description of the choice set.^12^ As research assistants led participants through the task, they would describe each option within a choice set to participants and point on the tablet to the attribute level they were describing. We selected KoBoToolbox, a freely available data collection application suited for rural settings because it allows for offline data collection. This is an important feature in settings with variable internet access. Finally, we aimed to make the appearance of each choice set easy to understand and follow. Choice sets were created in a tabular format to clearly depict the attributes and their corresponding attribute levels for each facility. We also incorporated images representing each attribute to remind respondents of the definition of that attribute (Figure 3). Images are also thought to help reduce the burden of the decision-making task by making it easier to summarize each presented option.^12^ These images were designed by Makerere Medical Illustrations after consultation with the lead author. The team of researchers then reviewed these images to ensure that they accurately represented each attribute. Self-administration of the DCE is an alternative administration approach researchers can consider. To support this approach in a low literacy population, a voice recording would need to be matched to the instructions, each attribute, and each attribute level for each unique choice set. Self-administration might be preferable in situations wherein research staff numbers are limited relative to the number of required participants.

Lastly, we pre-tested the tool by administering it to 7 randomly selected respondents from our target population.^9^ Pre-testing participants were presented 8 choice sets and asked to freely express their rationale for making each selection. This feedback revealed that respondents understood the tasks being asked of them and could complete all choice sets without significant cognitive burden. If the reported cognitive burden was too high during pre-testing, the tool would have required alteration, such as reducing the number of attributes or choice sets.

We have presented a detailed process guide for health researchers to reference in developing rational, locally-relevant DCEs for RLS. This guide describes the challenges we faced and decisions we made in the development of our DCE. We hope this guide will help increase the global use of DCEs as an informative tool to prioritize intervention design in RLS.

## Data Availability

The data that support the findings of this study are available from the corresponding author upon reasonable request.

## Notes

### Competing Interest Statement

The authors have declared no competing interest.

### Funding Statement

This work was supported by the Downs International Health Student Travel Fellowship and Yale Institute of Global Health Hecht Global Health Faculty Network Award. Dr. Hsieh is supported by the National Institute of Health/Fogarty International Center K01TW009995.

### Author Declarations

IRB approval was granted by the Yale University IRB and the Makerere University School of Medicine Research Ethics Committee.

